# Elevated Angiopoietin-2 inhibits thrombomodulin-mediated anticoagulation in critically ill COVID-19 patients

**DOI:** 10.1101/2021.01.13.21249429

**Authors:** Michael Hultström, Karin Fromell, Anders Larsson, Susan E Quaggin, Christer Betsholtz, Robert Frithiof, Miklos Lipcsey, Marie Jeansson

**Author notes:** **Corresponding author:** Marie Jeansson. **Competing interests:** The authors have declared that no conflict of interest exists.

## Abstract

Several studies suggest that hypercoagulation and endothelial dysfunction play central roles in severe forms of COVID-19 infections. We hypothesized that the high levels of the inflammatory cytokine Angiopoietin-2 (ANGPT2) reported in hospitalized COVID-19 patients might promote hypercoagulation through ANGPT2 binding to thrombomodulin with resulting inhibition of thrombin/thrombomodulin-mediated physiological anticoagulation. Plasma was collected from critically ill COVID-19 patients treated in the intensive care unit (ICU) at Uppsala University Hospital and ANGPT2 was measured at admission (61 patients) and after ten days (40 patients). ANGPT2 levels were compared with biochemical parameters, clinical outcome, and survival. We found that ANGPT2 levels were increased in COVID-19 patients in correlation with disease severity, hypercoagulation, and mortality. To test causality, we administered ANGPT2 to wildtype mice and found that it shortened bleeding time in a tail injury model. In further support of a role for ANGPT2 in physiological coagulation, bleeding time was increased in endothelial-specific *Angpt2* knockout mice. Using *in vitro* assays, we found that ANGPT2 inhibited thrombomodulin-mediated anticoagulation and protein C activation in human donor plasma. Our data reveal a novel mechanism for ANGPT2 in hypercoagulation and suggest that Angiopoietin-2 inhibition may be tested in the treatment of hypercoagulation in severe COVID-19 infection.

## Introduction

SARS-CoV-2 infection may be paucisymptomatic or lead to coronavirus disease-2019 (COVID-19), which has a wide range of symptoms and may cause severe illness, in particular in individuals with other cardiovascular risk factors (*1*). Thrombotic and thromboembolic disease have emerged as major COVID-19 complication despite routine thrombosis prophylaxis being standard of care (*2-5*). Microthrombosis has been suggested to contribute to both respiratory failure and neurological complications (*6, 7*), and activation of the coagulation system indicates a poor prognosis among COVID-19 patients in intensive care (*1, 7-9*).

Angiopoietin-2 (ANGPT2) is an inflammatory cytokine, the circulatory level of which correlates with adverse outcomes in several critical care syndromes, including acute respiratory disease syndrome (ARDS) and sepsis (reviewed in (*10*)). Elevated plasma ANGPT2 is a strong predictor of death in infection-mediated ARDS independent of the infectious agent (*11*), and elevated plasma ANGPT2 is further associated with disseminated intravascular coagulation (DIC) in conjunction with sepsis (*12*). In COVID-19, recent data show that the ANGPT2 level is a good predictor of ICU admission (*13*) and correlates with the severity of disease (*14, 15*).

ANGPT2 exerts its effects through different molecular mechanisms, the most well-studied being inhibition of Tie2 receptor signaling. This causes destabilization of the endothelium in most vascular beds and promotes inflammation, vascular leakage, impairment of the endothelial glycocalyx, and activation of α5β1 integrin signaling (*16-26*). The administration of Tie2 activating agents confers vascular protection and reduced mortality in experimental models of sepsis (*12, 27, 28*). Moreover, the recent discovery that ANGPT2 binds thrombomodulin (*29*) suggest that ANGPT2 may have additional and direct effects on the coagulation system. Thrombomodulin is constitutively expressed on the luminal surface of endothelial cells, where it is an important member of the intrinsic anticoagulant pathway and also an anti-inflammatory agent (*30*). Thrombomodulin inhibits the procoagulant functions of thrombin by binding and inhibiting its interaction with procoagulant substrates and instead promoting thrombin-catalyzed activation of protein C (APC) (*31*). Endothelial-specific knockout of thrombomodulin in mice disrupts APC formation and causes lethal thrombus formation (*32*), highlighting the potency of this pathway.

We hypothesized that the increased plasma levels of ANGPT2 observed in COVID-19 patients could contribute to hypercoagulation by inhibition of thrombomodulin-mediated activation of protein C. To investigate this, we measured plasma ANGPT2 and coagulation parameters in relation to clinical outcome in a cohort of critically ill COVID-19 patients and healthy blood donor controls. We further utilized experimental animals and *in vitro* assays to investigate if ANGPT2 could inhibit thrombomodulin-mediated anticoagulation and activation of protein C.

## Results

### COVID-19 patients admitted to the ICU

The study included a cohort of 61 patients admitted to the ICU at Uppsala University Hospital due to SARS-CoV-2 infection. Plasma samples were collected 1-4 days after admission and after 10-14 days for 40 patients with extended length of stay at the ICU. Plasma from 40 blood donors for the hospital blood bank were used as controls to generate normal ranges for ANGPT2, ANGPT1, VWF, and ADAMTS13. Patient demographic characteristics are summarized in Table 1.

**Table 1.** Patient demographic characteristic and comorbidities

|  | **All patients**<br>(n=61) |
| --- | --- |
| Women, n (%) | 12 (20) |
| Age, years | 60.9 (56.7-65.4) |
| Body weight, kg | 88.6 (83.5-93.9) |
| BMI, kg/m<sup>2</sup> | 30.1 (28.5-31.8) |
| COVID-19 day on ICU arrival | 9.2 (8.3-10.3) |
| Invasive ventilation | 49 (80) |
| 90-day mortality | 36 (59) |
| **Comorbidities, n (%)** |  |
| Pulmonary disease | 18 (30) |
| Hypertension | 38 (62) |
| Heart failure | 4 (7) |
| Peripheral vessel disease | 17 (28) |
| Previous thromboembolic event | 8 (13) |
| Diabetes | 21 (34) |
| Malignancy | 6 (10) |
| ≥2 risk factors | 32 (26) |
| **Clinical features at arrival** |  |
| ARDS, n (%) | 20 (100) |
| SAPS3 score | 53 (51-56) |

Data are expressed as n (%) or geometric mean ± 95% CI. *BMI* body mass index; *ICU* intensive care unit; *ARDS* acute respiratory distress syndrome; *SAPS3* simplified acute physiology score.

### ANGPT2 is elevated in critically ill COVID-19 patients

Assessment of plasma ANGPT2 concentrations revealed that ANGPT2 was significantly elevated (p<0.0001) in patients at ICU admission compared to controls (Table 2). At 10-14 days after admission, ANGPT2 was further increased (p<0.01) (Table 2). ANGPT2 levels were further analyzed with regards to survival. ANGPT2 was significantly higher, both at admission and at 10-14 days, in non-recovering patients compared to recovering patients (p<0.01 and p<0.001, respectively) (Figure 1A). We then further analyzed ANGPT2 and survival by using Kaplan-Meier plots (Figure 1B, C). Using the optimal cut-off value of 8.3 ng/ml for ANGPT2 (please refer to Statistics for details), we demonstrated that ANGPT2 level at admission was strongly predictive of death (p<0.0001) but also at the later time point (p<0.0001) in the studied cohort. Plasma concentrations of the Tie2 agonist ANGPT1 were minimally affected (Figure 1D). Other clinical and measured parameters of the patients during their ICU stay are summarized in Table 2.

**Table 2.** Clinical parameters of patients during ICU stay.

|  | Day 1-4 | n | Day 10-14 | n | p-value<sup>A</sup> | Reference range | p-value<sup>B</sup> |
| --- | --- | --- | --- | --- | --- | --- | --- |
| ANGPT2 (ng/ml) | 3.1 (2.6-3.8) | 61 | 4.8 (4.0-5.8) | 40 | ** | 1.2 (1.9-1.4)<sup>C</sup> | **** |
| ANGPT1 (ng/ml) | 6.2 (4.9-7.8) | 61 | 5.2 (4.4-6.3) | 40 | ns | 9.9 (7.3-13.4)<sup>C</sup> | ns |
| VWF (IU/ml) | 3.4 (3.2-3.7) | 61 | 3.6 (3.2-4.0) | 40 | ns | 1.5 (1.4-1.7)<sup>C</sup> | **** |
| ADAMTS13 (ng/ml) | 887 (789-996) | 61 | 640 (524-782) | 40 | ** | 930 (844-1025)<sup>C</sup> | ns |
| Platelets (x10<sup>9</sup>/l) | 231 (208-258) | 58 | 426 (353-514) | 37 | **** | 150-300 | ns |
| D-dimer (mg/l) | 2.27 (1.70-3.03) | 60 | 2.49 (1.85-3.35) | 34 | ns | <0.50 | **** |
| CRP (mg/l) | 192 (164-224) | 59 | 65 (45-95) | 38 | **** | <5 | **** |
| SOFA score | 6 (6-7) | 60 | 6 (5-7) | 40 | ns | 0 | **** |
| PaO<sub>2</sub>/FiO<sub>2</sub> | 18.6 (17.0-20.3) | 57 | 23.2 (20.9-25.7) | 37 | ** | >60 | **** |
| Ferritin (µg/l) | 1377 (1033-1837) | 56 | 1417 (979-2068) | 36 | ns | 25-310 | **** |
| Lactate (mmol/l) | 1.3 (1.2-1.4) | 59 | 1.2 (1.1-1.4) | 38 | ns | 0.8-2.0 | ns |
| TNFα (ng/l) | 66.7 (56.4-79.0) | 40 |  |  |  | <8.0 | **** |
| IL6 (ng/l) | 156 (120-204) | 35 |  |  |  | <7.0 | **** |
| Fibrinogen (g/l) | 8.4 (7.6-9.3) | 15 |  |  |  | 2.0-4.2 | **** |
| TEG |  |  |  |  |  |  |  |
| R(min) | 8.7 (7.5-10.0) | 30 | 12.3 (8.6-17.5) | 22 | ns | 4.6-9.1 | ns |
| R<sub>KCH</sub> (min) | 7.3 (6.8-8.0) | 30 | 9.2 (8.0-10.6) | 22 | ** | 4.3-8.3 | ns |
| Angle (deg) | 77.1 (75.8-78.4) | 30 | 76.5 (74.2-78.9) | 22 | ns | 64-77 | ns |
| MA (mm) | 68.8 (67.2-70.4) | 30 | 73.6 (72.1-75.1) | 22 | **** | 52-69 | ns |

Data are expressed as geometric mean ± 95% CI. CRP C-reactive protein; PaO<sub>2</sub>/FiO<sub>2</sub> the ratio of arterial oxygen partial pressure (PaO<sub>2</sub> in mmHg) to fractional inspired oxygen; TEG thromboelastography, R reaction time; KCH kaolin with heparinase; MA maximum amplitude.
<sup>A</sup>p-values from unpaired t-test on log transformed data comparing patients at day 1-4 and patients at day 10-14.
<sup>B</sup>p-values from Fisher's exact test comparing patients at day 1-4 with reference range.
<sup>C</sup>measured in healthy controls in the study.

**Figure 1.**
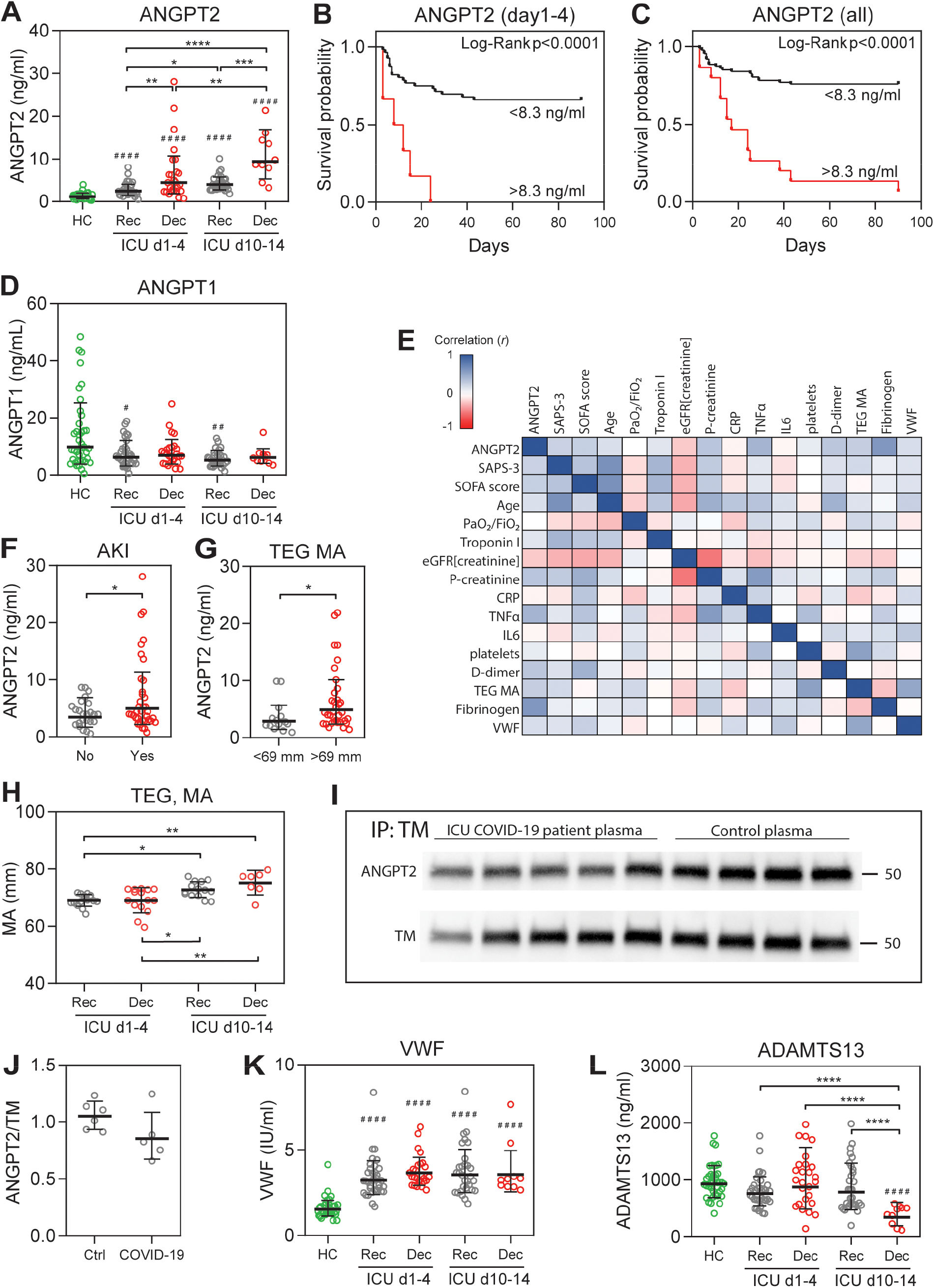
Plasma ANGPT2 is increased in critically ill COVID-19 patients and correlates to mortality and hypercoagulation. (A) Plasma ANGPT2 concentrations for healthy controls (HC), recovered (Rec) and deceased (Dec) patients at day 1-4 and day 10-14 after admission. (B, C) Kaplan Meier plots with Log-rank test show that ANGPT2 levels are higher in non-recovering patients both at admission and when including all time-points and can predict mortality. (D) Plasma ANGPT1 was minimally affected. (E) Clinical data and measured parameters were analyzed by Pearson correlation test (Spearman for SAPS-2 and SOFA score) and presented in a heatmap for correlation (blue) or inverse correlation (red). A summary of ANGPT2 comparisons is presented in Table 3, and all data in Suppl. Table 1. (F) ANGPT2 is higher in patients with acute kidney injury (AKI) compared to non-AKI patients. (G) ANGPT2 is higher in patients with MA >69. (H) Thromboelastography (TEG) measurements for MA, other TEG measurements can be seen in Table 2. (I, J) Immunoprecipitation and quantification of thrombomodulin (TM) from plasma in controls and COVID-19 patients show binding of ANGPT2. Full uncut blots available in Suppl Fig 1. (K, L) Plasma concentrations for VWF and ADAMTS13, respectively. Data shown as shown as geometric mean ± 95% CI (A, D, F-H, J-L).^# # # #^ p<0.0001 vs. HC, ^*^p<0.05, ^**^p<0.01, ^***^p<0.001, ^****^p<0.0001. p-value from one-way ANOVA with Bonferroni post hoc test (A, D, H, K, L), and t-test (F, G, J) on log transformed data. *TEG* thromboelastography; *MA* maximal amplitude

Further analysis showed that ANGPT2 correlated with several markers of disease severity, shown in a heatmap representing the degree of correlation (*r*) (Figure 1E), with values and significance in Suppl. Table 1. The simplified acute physiology (SAPS-3) score (probability of death) correlated significantly with ANGPT2 at ICU admission (p<0.05) (Table 3). Sequential organ failure assessment (SOFA) score was calculated for the same days as collection of plasma samples. The SOFA score represents six organ systems, were each organ system is assigned a point value from 0 (normal) to 4 (high degree of dysfunction/failure) (*33*). ANGPT2 and SOFA score were significantly correlated (p<0.01) (Table 3). In line with this, we found that COVID-19 patients with acute kidney injury (AKI) had significantly (p<0.05) higher ANGPT2 levels (Figure 1F) and that ANGPT2 inversely correlated (p<0.0001) with eGFR[creatinine] (Table 3). In contrast, we found no association of ANGPT2 and troponin I (myocardial injury) or pulmonary function (PaO_2_/FiO_2_) (Table 3). The inflammatory markers CRP, TNFα, IL6, and ferritin were all significantly (p<0.0001) elevated in COVID-19 patients at admission (Table 2). TNFa showed a significant (p<0.01) correlation with ANGPT2 (Table 3). The other inflammatory markers did not correlate with ANGPT2 and neither did lactate.

**Table 3.** Correlation of ANGPT2 and other parameters in the cohort.

| ANGPT2 vs. | r | p | n |
| --- | --- | --- | --- |
| SAPS-3 | 0.270 | * | 58 |
| SOFA score | 0.286 | ** | 100 |
| Age | 0.183 | ns | 61 |
| PaO<sub>2</sub>/FiO<sub>2</sub> | -0.007 | ns | 94 |
| eGFR<sup>[creat]</sup> | -0.352 | **** | 97 |
| Troponin I | 0.152 | ns | 94 |
| CRP | -0.078 | ns | 95 |
| TNFα | 0.442 | ** | 40 |
| IL6 | -0.063 | ns | 41 |
| Platelet count | 0.118 | ns | 95 |
| D-dimer | 0.330 | * | 94 |
| Fibrinogen | 0.609 | * | 15 |
| TEG MA | 0.287 | * | 49 |
| VWF | 0.200 | * | 101 |

p-value from Pearson's correlation test for all parameters except SAPS-3, SOFA score, and TEG MA where Spearman's correlation test was used.

### Hypercoagulation in critically ill COVID-19 patients

To investigate if ANGPT2 correlated with hypercoagulation in these patients, we assessed several markers of the coagulation system summarized in Table 2. Of note, all patients received prophylactic anticoagulation therapy with Dalteparin sodium during the ICU stay (*5*). Platelet counts were significantly (p<0.0001) increased in patients at the late timepoint compared to the reference interval and the early timepoint (Table 2) and showed significant (p<0.05) correlation with ANGPT2 (Table 3). D-dimer levels were increased (p<0.0001) in all patients and correlated significantly (p<0.05) with ANGPT2 (Table 2, 3). Fibrinogen had similar pattern and correlated (p<0.05) with ANGPT2 (Table 2, 3). Thromboelastography (TEG) was performed on some patients during their ICU stay. TEG maximal amplitude (MA), representing thrombus strength, was significantly increased over time in the cohort (p<0.0001) (Figure 1H, Table 2). The patients with an MA value >69 mm had significantly (p<0.05) higher ANGPT2 levels (Figure 1G) and there was a significant (p<0.05) correlation between MA and ANGPT2 (Table 3). In contrast, reaction time (R) increased over time in the cohort (p<0.01) (Table 2). We did not find differences in other TEG parameters (Table 2). Although we were unable to assess more direct measures of thrombomodulin effects (like activated protein C), immunoprecipitation of circulating thrombomodulin from patient and control plasma showed binding of ANGPT2 (Figure 1I, J).

In addition to ANGPT2, other factors are likely to play additional roles for hypercoagulation in COVID-19. Von Willebrand factor (VWF) is stored and released from platelets and endothelial Wiebel Palade bodies (*34*). Because VWF and ANGPT2 have been reported to be stored in the same endothelial vesicles (*35*), we assessed plasma levels of VWF. VWF was significantly (p<0.0001) elevated in all COVID-19 patients compared to healthy controls and correlated (p<0.05) to ANGPT2 (Figure 1K, Table 2, 3). ADAMTS13 is a metalloprotease produced by the liver that degrades large VWF multimers, thereby decreasing VWF’s procoagulation properties (*36*). We found that ADAMTS13 was significantly (p<0.0001) decreased at the late timepoint in patients later dying from COVID-19 compared to all other patients, suggesting an increased consumption of ADAMTS13 (Figure 1L). VWF correlated with ANGPT2, SOFA score, eGFR[creatinine], P-creatinine, platelet count, and TEG MA, but not with SAPS-3 and could not predict mortality at any time-point (Suppl. Table 1). Taken together, our data strongly suggest a link between ANGPT2 and the coagulation system.

### ANGPT2 administration decreases tail bleeding time in mice

To expand on the *in vitro* studies performed by Daly et al (*29*) showing that ANGPT2 binds thrombomodulin, we performed *in vivo* studies in mice to evaluate the effect of ANGPT2 on bleeding time. We employed the tail bleeding model, which has been used extensively to evaluate the coagulation system in mice (*37*). Recombinant His-tagged human ANGPT2, ANGPT1, or IgG as controls, were injected 15 minutes before tail bleeding. In these experiments, 250 µg/kg of ANGPT2 resulted in significantly (p<0.01) reduced bleeding time, whereas ANGPT1 did not differ from the IgG control (Figure 2A). The resulting plasma concentrations after ANGPT2 injection correlated roughly with the injected dose (Figure 2B). To examine if ANGPT2 bound directly to thrombomodulin, we performed experiments on lung tissue after injection of 250 ug/kg ANGPT2, ANGPT1 or IgG. Thrombomodulin was immunoprecipitated, and the signals for His-tag and total thrombomodulin were evaluated. ANGPT2 showed significantly (p<0.01) more binding to thrombomodulin compared to IgG or ANGPT1 (Figure 2C, D).

**Figure 2.**
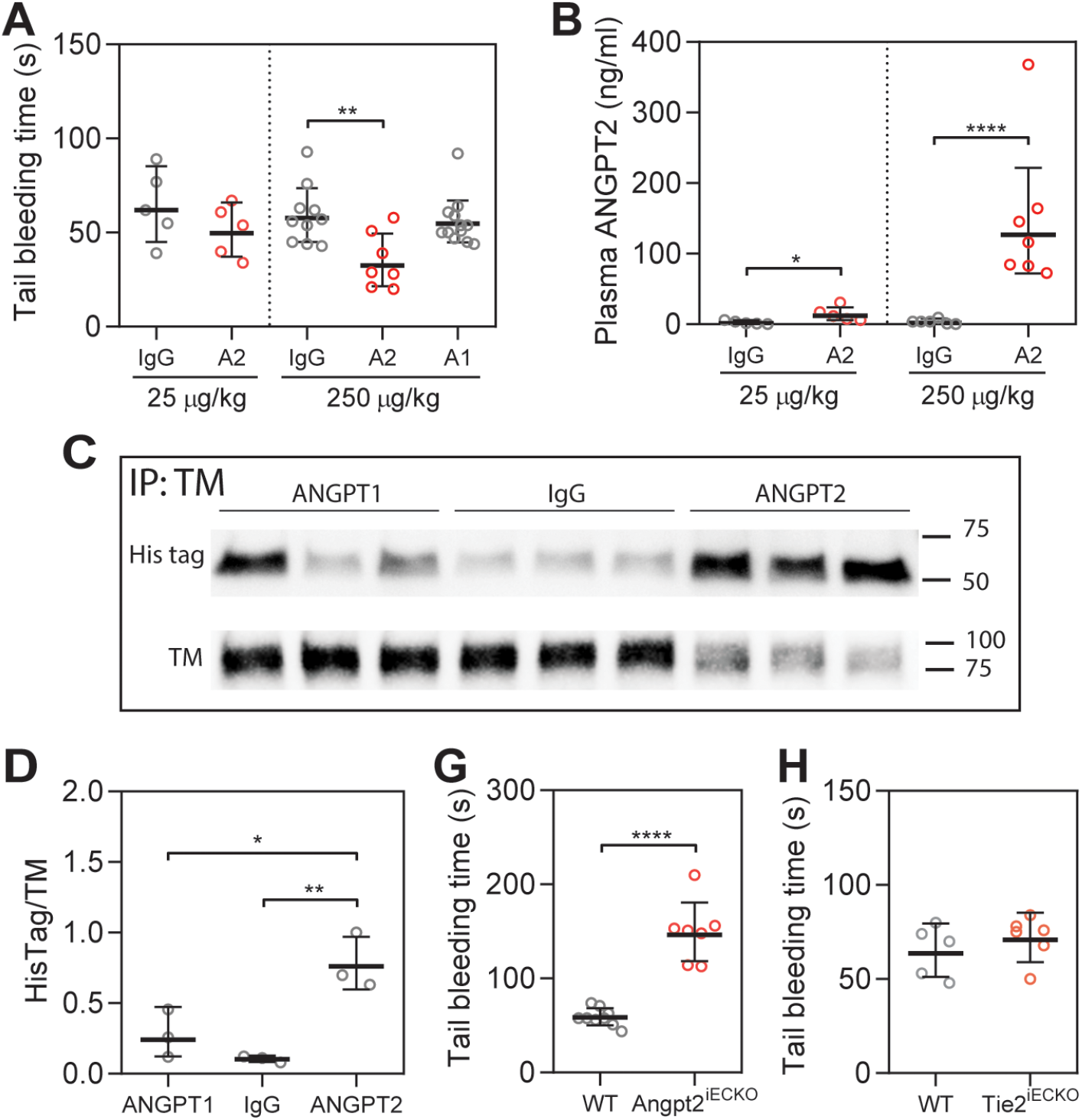
ANGPT2 bind thrombomodulin and shortens tail bleeding time in mice. (A) Tail bleeding time 15 min after i.p. injection of His tagged ANGPT2 (A2), and ANGPT1 (A1) at indicated doses. Albumin or IgG was injected in controls. (B) Plasma concentrations of ANGPT2 after i.p injection. (C, D) Immunoprecipitation and quantification of thrombomodulin (TM) from lung lysates in injected mice with blotting for His tag and TM. (G, H) Tail bleeding time in Angpt2^iECKO^ mice and Tie2^iECKO^ mice, respectively. Data presented as geometric mean ± 95% CI. ^*^p<0.05, ^**^p<0.01, ^****^p<0.0001. p-value from one-way ANOVA with Bonferroni post hoc test (A, B, D), and t-test (G, H) on log transformed data.

In contrast, mice with an induced endothelial-specific knockout of *Angpt2* showed significantly (p<0.0001) increased bleeding time (Figure 2G). ANGPT2’s effect on tail bleeding time was Tie2 independent, as endothelial-specific *Tie2* knockouts showed no differences in bleeding time (Figure 2H). Previous studies showed lack of a difference in tail bleeding time in *Angpt1* knockout mice (*29*).

### ANGPT2 inhibits thrombomodulin mediated anticoagulation and activation of protein C

To further study the effects of ANGPT2 on coagulation, we utilized thromboelastography (TEG) on human plasma supplemented with thrombomodulin and ANGPT2. A TEG curve from one of the high responder donors can be seen in Figure 3A with measured parameters indicated. As expected through its negative regulation on the coagulation system, thrombomodulin significantly (p<0.001) increased the time for coagulation to start (reaction time – TEG R) (Figure 3B, Table 4). This effect was significantly inhibited by ANGPT2 (p<0.01). Trends towards a thrombomodulin-induced decrease in thrombus strength (maximal amplitude – TEG MA) and its inhibition by ANGPT2 were also observed, although these effects were not statistically significant (Figure 3C). It should be noted that plasma from all donors responded to thrombomodulin and ANGPT2 but to a variable degree. ANGPT2 on its own did not affect TEG parameters (Figure 3B, C, Table 4).

**Table 4.** TEG data from in vitro experiments

|  | Control | TM | TM+ANGPT2 | ANGPT2 |
| --- | --- | --- | --- | --- |
| R(min) | 8.5 (6.8-10.6) | 10.0 (7.4-13.6)* | 9.2 (7.0-11.9) | 8.0 (3.8-16.6) |
| Angle (deg) | 65.3 (60.7-70.1) | 54.6 (42.1-70.9) | 59.5 (50.7-69.7) | 67.8 (59.4-77.4) |
| MA (mm) | 44.8 (36.3-55.2) | 39.5 (28.2-55.2) | 43.5 (34.2-55.3) | 45.3 (24.8-82.8) |

Data is presented as geometric mean ± 95% CI. *R* reaction time; *Angle* rate of clot formation; *MA* maximal amplitude, *TM* thrombomodulin (1000 ng/ml), *ANGPT2* angiopoietin-2 (1000 ng/ml). Statistical comparison was done with repeated measures one-way ANOVA on log transformed data.

**Figure 3.**
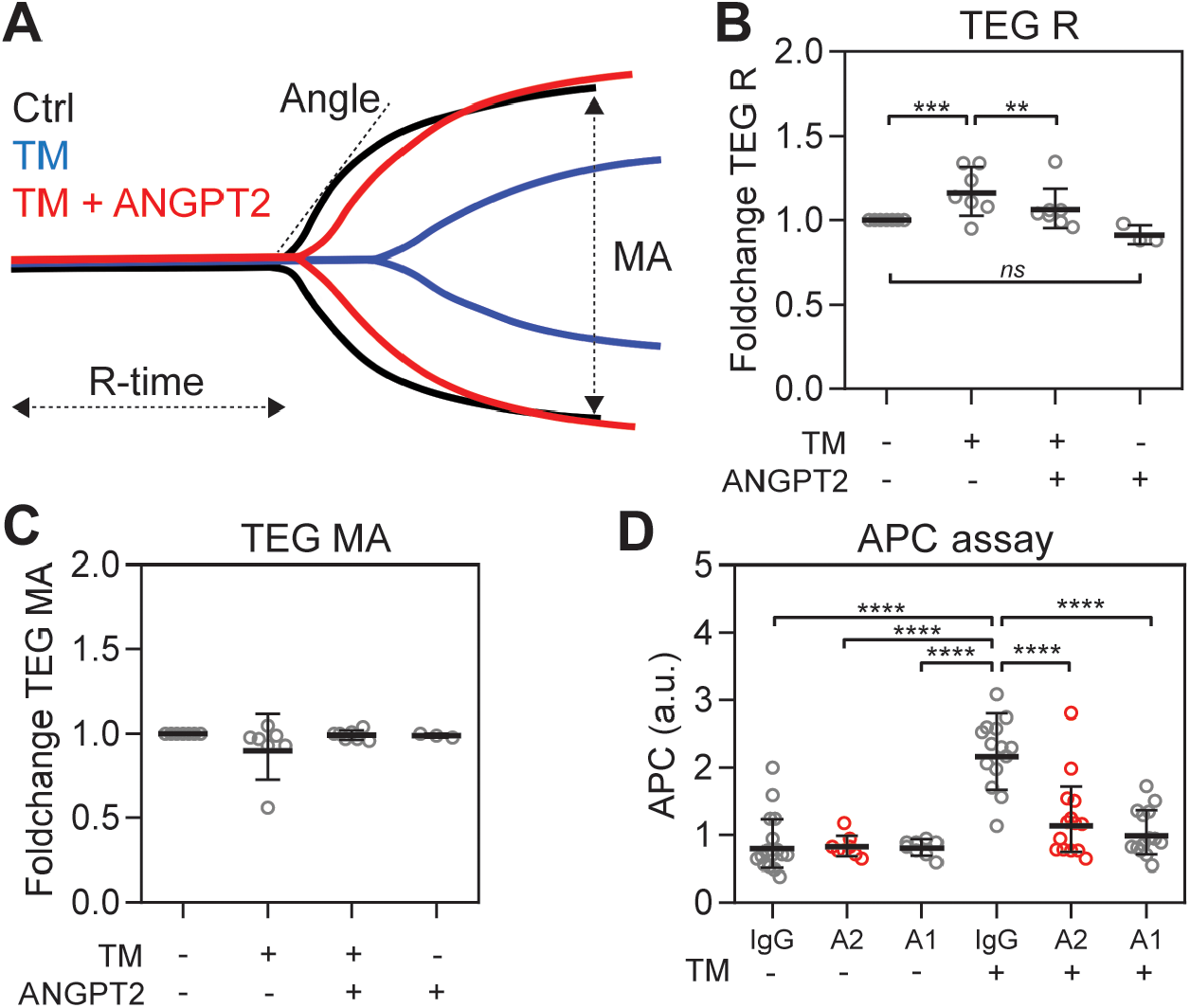
ANGPT2 inhibits thrombomodulin mediated anticoagulation and protein C activation. TEG curve from one of the donors showing thrombomodulin (TM) dependent increase in reaction time (R) and decreased maximal amplitude (MA), which is inhibited by ANGPT2 (A). TEG analysis of individual donor blood with addition of 1000 ng/ml thrombomodulin (TM) and 1000 ng/ml ANGPT2. Foldchange graphs from TEG analysis for R (B) and MA (C). Group results from these experiments can be found in Table 4. Thrombomodulin dependent formation of APC can be inhibited with ANGPT2 or ANGPT1 (D). Data presented as geometric mean ± 95% CI. ^**^p<0.01, ^***^p<0.001, ^****^p<0.0001. p-value from one-way ANOVA with Bonferroni post hoc test on log transformed data. *TEG* thromboelastography

Next, we investigated ANGPT2’s effect on activation of protein C in human plasma *in vitro*. Human plasma was incubated with thrombomodulin with ANGPT2, ANGPT1, and control IgG to study thrombin/thrombomodulin mediated activation of protein C. Activated protein C was measured with chromogenic APC substrate. As expected, the addition of thrombomodulin significantly (p<0.0001) increased activation of protein C compared to IgG, ANGPT2, and ANGPT1 alone (Figure 3D). Both ANGPT2 and ANGPT1 significantly (p<0.0001) reduced thrombomodulin-mediated activation of protein C (Figure 3D).

## Discussion

Our comprehensive translational approach, comprising analysis of plasma and clinical features in critically ill COVID-19 patients together with mechanistic studies in mice and *in vitro*, suggests a novel role for ANGPT2 in COVID-19-associated hypercoagulation. We found that elevated ANGPT2 correlated with markers of the coagulation system in plasma in COVID-19 patients, with the highest levels in patients that subsequently died from the disease. Using mice, we further found a procoagulant effect of administered ANGPT2 and an anti-coagulant effect of genetic inactivation of the *Angpt2* gene. *In vitro* experiments with human plasma showed that ANGPT2 inhibited thrombomodulin-mediated anticoagulation and protein C activation. Taken together, our data suggest that elevated ANGPT2 might have an important pathogenic role in critically ill COVID-19 patients, and potentially also in other diseases with hypercoagulation. Our suggested function of ANGPT2 in hypercoagulation is summarized in Figure 4.

**Figure 4.**
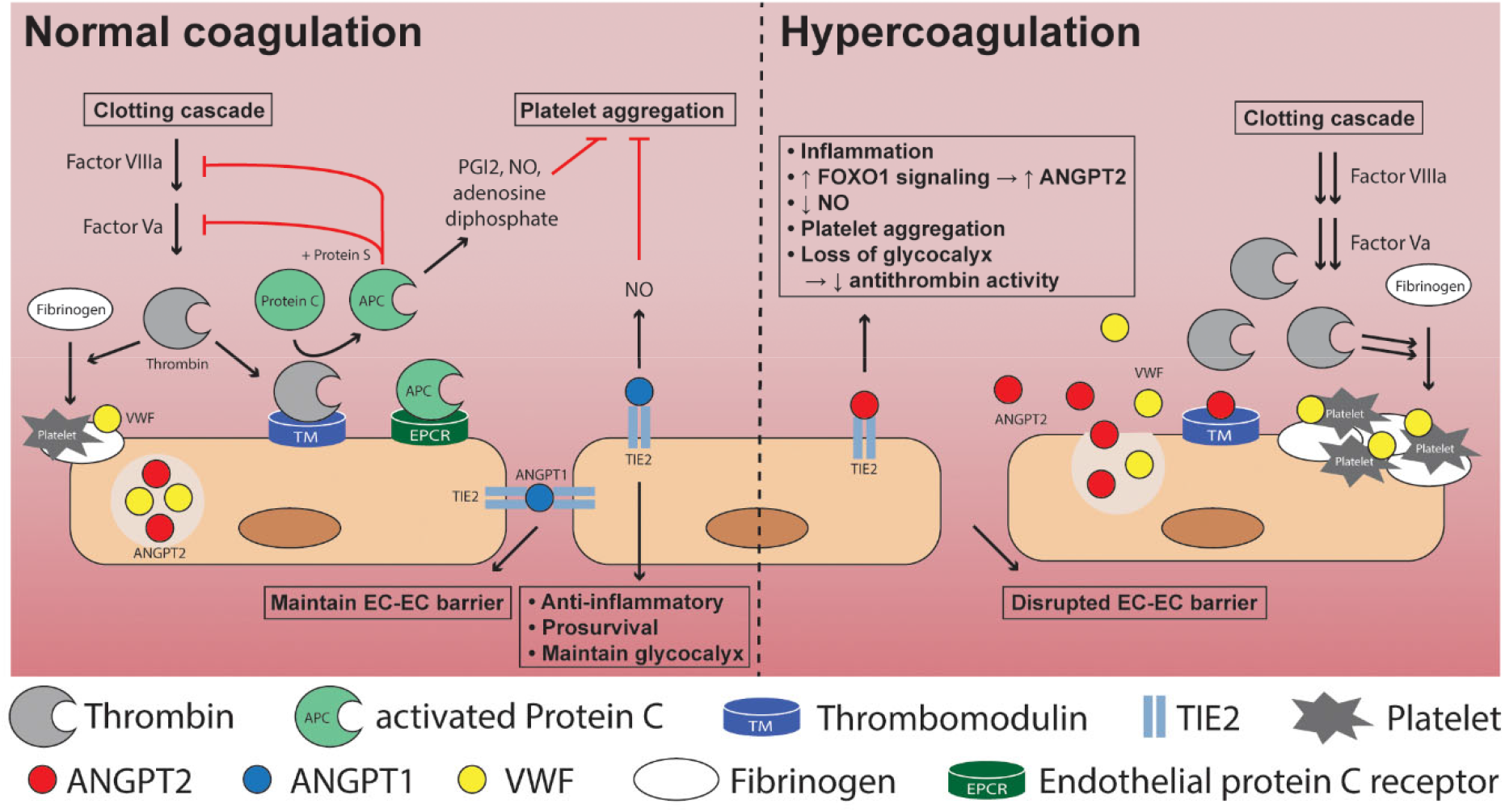
A schematic overview of Angiopoietin signaling in normal coagulation and in hypercoagulation with high ANGPT2. In addition to ANGPT2 inhibition of thrombomodulin mediated anticoagulation several other endothelial functions are disturbed by high ANGPT2.

Circulating ANGPT2 levels correlated with severity of disease, hypercoagulation, and mortality in the studied cohort. Our results for ANGPT2 are in line with recently published data in COVID-19 patients (*13-15, 38*). The strength of our study is that we analyzed two different timepoints in these patients, at admission to the ICU and 10-14 days after admission. This revealed that ANGPT2 is increased over time in all patients in the ICU, and that non-survivors have an even further increase. We find that ANGPT2 levels at admission can significantly predict mortality and correlates to the ICU indices SAPS-3 (probability of death). In addition, we find that ANGPT2 correlates significantly to indices of organ failure (SOFA score) including kidney injury (AKI, eGFR[creatinine]), but not myocardial injury (troponin I), nor pulmonary function (PaO_2_/FiO_2_).

To investigate if ANGPT2 levels correlated with hypercoagulation in these patients, we assessed several markers of the coagulation system. Ideally, we would have wanted to analyze a more specific target for our hypothesis i.e. activated protein C, but this was unsuccessful. Instead, we studied clinical available markers of the coagulation system activation including D-dimer, fibrinogen, platelet count, and TEG. All these markers indicate a hypercoagulative state in these patients that worsens over time. More importantly, we find that ANGPT2 levels correlate with all of them i.e. D-dimer, fibrinogen, platelet count, and maximal amplitude (MA) from TEG. The risk of developing venous thromboembolism in orthopedic trauma patients in known to increase when MA ≥ 65 mm, and further doubles when MA ≥ 72 mm (*39*). In this study, patients with MA >69 mm (upper reference interval) had significantly higher ANGPT2. Increased MA has previously been reported in critically ill COVID-19 patients (*5, 40, 41*). Recently, thrombin-antithrombin complexes and factor VIIIa were shown to be significant upregulated in critically ill ICU-contained COVID-19 patients compared to hospitalized non-ICU COVID-19 patients (*42*). D-dimer is widely reported to be increased in COVID-19 patients.

VWF was significantly increased in all ICU patients compared to healthy controls, but in contrast to ANGPT2, we found no differences in VWF levels between the ICU patient groups. This contrasts to a recent study reporting a correlation between VWF and mortality in ICU COVID-19 patients (*42*). Nevertheless, because of the suggested co-storage of ANGPT2 and VWF in endothelial Weibel Palade bodies (*35*), we wanted to pursue the VWF track, and therefore investigated ADAMTS13, a liver-derived plasma metalloprotease that degrades large VWF multimers and thereby decreases VWF’s pro-hemostatic properties (*36*). Previously, decreased concentrations of ADAMTS13 have been shown to correlate with mortality in COVID-19 and septic shock patients (*15, 43*). Interestingly, ADAMTS13 levels decreased over time in the ICU in non-recovering patients in our cohort. It is plausible that this progressive decrease reflects increased consumption rather than lowered production of ADAMTS13.

Endothelial dysfunction can be mediated by ANGPT2 through its binding to Tie2 (*44*) and integrin β1α5 (*26*) (Figure 4). More recently, Daly *et al* reported that ANGPT2 can bind thrombomodulin and inhibit thrombomodulin-mediated anticoagulation *in vitro* (*29*).

Thrombin coupled to thrombomodulin converts protein C to activated protein C (APC), an endogenous protein that promotes fibrinolysis and inhibits thrombosis and inflammation (*30*) (Figure 4). ANGPT2 (and ANGPT1) binding to thrombomodulin inhibits its binding to thrombin and subsequent activation of protein C (*29*). APC formation may also be impaired because of down-regulation or shedding of thrombomodulin induced by inflammatory cytokines (*45*), and increased circulating levels of thrombomodulin have been reported in COVID-19 patients which correlated with mortality (*15, 42*). We utilized circulating thrombomodulin in immunoprecipitation experiments from COVID-19 patient and control plasma. We found that circulating thrombomodulin carried bound ANGPT2, showing that this interaction is indeed happening *in vivo* in both controls and patients (Figure 1). We did not find any significant difference in the ratio between ANGPT2/thrombomodulin pulled down together with thrombomodulin. We speculate that under normal conditions there is a low steady state of thrombomodulin bound ANGPT2, which increases in COVID-19 with high ANGPT2 levels and increased shedding of thrombomodulin. More studies are needed to investigate this further. While reduced levels of APC are found in a majority of patients in sepsis and are associated with increased risk of death (*46-49*), a recent study did not find changes of APC in critically ill COVID-19 patients (*42*).

To further investigate if ANGPT2 can directly affect the coagulation system *in vivo*, we performed several experiments in mice. One simple but highly relevant experiment to evaluate coagulation is tail bleeding time (*37*). In these experiments, recombinant ANGPT2 and ANGPT1 were injected before the measurement of tail bleeding time. These experiments showed that ANGPT2, but not ANGPT1, could decrease bleeding time *in vivo*. Immunoprecipitation of thrombomodulin from lung tissue showed binding of ANGPT2 (Figure 2). In contrast, mice with endothelial-specific deletion of *Angpt2* displayed longer bleeding times. Furthermore, endothelial-specific deletion of *Tie2* did not change bleeding time, excluding a Tie2-dependent mechanism in our model. Bleeding time in *Angpt1* knockout mice was reported to be normal in a previous study (*29*). In contrast, Higgins *et al*, reported that heterozygous *Tie2* knockout mice had an increased thrombotic response at the site of laser injury (*12*). As Tie2 signaling regulates the transcription of *Angpt2* (*50*), it is tempting to speculate that heterozygous *Tie2* knockout mice had more Angpt2 protein stored in endothelial secretory vesicles, and that increased local release of Angpt2 occurred upon laser injury. Regardless, it is evident that more studies are needed concerning this pathway and its implications for disease.

TEG was also used to study ANGPT2 inhibition of thrombomodulin-mediated anticoagulation in freshly collected plasma from healthy donors. All donor plasma had a thrombomodulin-mediated increase in reaction time that could be inhibited by ANGPT2 (Figure 3). The ability of ANGPT2 and ANGPT1 to inhibit thrombomodulin-mediated APC production was investigated in an APC assay with human plasma and a chromogenic APC substrate. In this assay, we found that both ANGPT2 and ANGPT1 could significantly reduce activation of protein C by approximately 50% (Figure 3), which agrees with data reported by Daly *et al* (*29*). Why is this effect elicited by ANGPT1 *in vitro*, but not *in vivo*? A possible answer is provided by Daly *et al*, who suggest that in the presence of both thrombomodulin and TIE2, ANGPT1 preferably binds TIE2, whereas ANGPT2 binds both thrombomodulin and TIE2 (*29*). Currently, we have no means of assessing the local concentrations of ANGPT2 and ANGPT1 *in vivo*. Endothelial expression of ANGPT2 varies extensively among vessel types and location and is upregulated in response to angiogenic and inflammatory activation. It is therefore possible that inhibition of thrombomodulin occurs only locally at sites of high ANGPT2 release. ANGPT1 on the other hand, is important for endothelial stabilization and anti-inflammatory properties through TIE2 signaling. In line with this, release of ANGPT1 in conjunction with platelet degranulation and resulting signaling through TIE2 have been shown to be important for endothelial barrier closure after neutrophil extravasation (*51*). Although ANGPT1 might inhibit thrombomodulin *in vitro*, this may not be a major concern *in vivo*, as most studies show unchanged or decreased ANGPT1 in disease, including the current study and a large sepsis study (*12*).

Our data suggest that inhibition of ANGPT2 may be explored as a therapeutic approach in COVID-19 and other diseases with hypercoagulation. A compound known as Trebananib (formerly AMG386) binds to both ANGPT2 and ANGPT1 and inhibits interaction with TIE2 (*52*). Trebananib has been tested in multiple clinical cancer trials and has been administered to a large number of patients (*53-55*), and to our knowledge, coagulation disorders have not been reported as adverse effects in these trials. While Trebananib might be an interesting compound to test in COVID-19 patients, its binding and inhibition of ANGPT1-TIE2 signaling may complicate matters, as ANGPT1-TIE2 signaling in known to protect the vasculature and decrease inflammation (*28*). Inhibiting ANGPT1 may therefore have adverse effects. There are currently many clinical trials registered with Angiopoietin-2 antibodies, several for the treatment of solid tumors. Critically ill COVID-19 patients may benefit from these antibodies. Another very interesting compound at the preclinical stage is ABTAA, a humanized <u>A</u>ngiopoietin-2 <u>B</u>inding and <u>T</u>ie2-Activating <u>A</u>ntibody (*28*). ABTAA binds and clusters ANGPT2, converting it into a TIE2-activating molecule while simultaneously decreasing free ANGPT2, which antagonizes TIE2 signaling. ABTAA treatment has shown promising results in experimental models of sepsis (*28*), however, coagulation was not evaluated. Another approach would be to interfere with exocytosis and translation of ANGPT2. Several compounds can stimulate exocytosis of ANGPT2 *in vitro*, including thrombin, histamine and TNFα (*21, 56*). We found a significant correlation between ANGPT2 and TNFα, and patients may benefit from anti-TNFα treatments (*57*). Studies have shown that serum concentrations of cytokines and acute phase proteins are downregulated after administration of anti-TNFα therapy in patients with rheumatoid arthritis, including IL-6, IL-1 receptor antagonist, serum amyloid A, haptoglobin, and fibrinogen (*58-60*). Markers of the coagulation system are also rapidly downregulated, with significant reductions in D-dimer and pro-thrombin fragments seen shortly after anti-TNFα therapy (*61*).

We acknowledge certain limitations in our study. First, this study was neither designed nor powered to test the performance of parameters for outcome prediction. However, our findings are plausible, hypothesis-generating, and clearly deserve validation in a larger cohort of patients. Second, most of the COVID-19 patients were male, an overrepresented sex among COVID-19 patients in intensive care at the time of our study, and inference of our results to female COVID-19 patients should therefore be made with caution. Importantly, experiments in mice and analyses on donor blood had representation of both sexes.

In conclusion, we show that ANGPT2 levels in critically ill COVID-19 patients correlate with severity of disease, hypercoagulation, and mortality. In addition, we provide novel *in vivo* evidence for a direct role for ANGPT2 in coagulation through binding to and inhibition of thrombomodulin-mediated anticoagulation. These findings suggest that inhibition of ANGPT2 might not only benefit critically ill COVID-19 patients but also other patients with hypercoagulation.

## Methods

### Study design and patients

The present study is part of a prospective single-center observational study at the ICU at Uppsala University Hospital. During the first wave between March 13 and August 14, 2020, 123 patients older than 18 years of age were included in the study. All patients were diagnosed with COVID-19 by positive reverse-transcription PCR from nasopharyngeal swabs. Samples from 61 patients were included in the study based on severity of disease and length of stay. 40 patients with extended length of stay >10 days were included with one sample at admission, and one sample after at least 10 days. In addition, 20 patients with severe disease but shorter length of stay were included with one sample at admission only. Apart from the customary ICU care and medications, all patients received thromboprophylaxis with dalteparin sodium at 100 IU/kg. Blood was sampled in 0.129 M trisodium citrate tubes (9NC BD Vacutainer, Becton Dickinson) and stored as plasma at -80°C until analysis.

Clinical data were recorded prospectively, including medical history, medications, physiological data, and date of death. The Simplified Acute Physiology Score 3 (SAPS-3) and Sequential Organ Failure Assessment (SOFA) score, circulatory support, and respiratory support data were collected as detailed in the Results section (Table 1, 2).

Healthy control plasma was collected after consent from forty consecutive adult blood donors (43% females, 43 (37-49) years old) visiting the Blood Central at Uppsala University Hospital on June 23, 2020. TEG was performed on freshly collected plasma from 7 donors (4 females/3 males, 30-50 years old). In addition, freshly collected plasma from 5 ICU hospitalized COVID-19 patients were used for immunoprecipitation of thrombomodulin and ANGPT2 blotting.

### Blood examinations

All routine lab tests were performed at the hospital’s clinical chemistry department. First blood sample was collected 1-4 days after admission, the second 10-14 days after admission. ELISA’s were used to measure plasma protein concentrations for Angiopoietin-2 (DANG20, R&D Systems), Angiopoietin-1 (DANG10, R&D Systems), von Willebrand factor (ab108918, Abcam), and ADAMTS13 (ab234559, Abcam) according to the manufacturer’s instructions. To investigate if binding of ANGPT2 to thrombomodulin occurred in patients, we utilized immunoprecipitation of circulating thrombomodulin in plasma and blotted for ANGPT2 (details below).

### Mice

Floxed *Angpt2* mice (*62*) and *Tie2* mice (*62*) were crossed to tamoxifen inducible *Cdh5*-Cre^ERT2^(*63*) mice to generate endothelial specific knockout of *Angpt2* (*Angpt2*^iECKO^) and *Tie2* (Tie2^*iECKO*^). Controls were littermate mice with wt/wt alleles for *Angpt2* (WT) and *Tie2* (WT). Mice were genotyped with primers for Angpt2 (for 5’-GGGAAACCTCAACACTCCAA and rev 5’-ACACCGGCCTCAAGACACAC, wt 222 bp, floxed 258 bp), Tie2 flox (for 5’-TCCTTGCCGCCAACTTGTAAAC and rev 5’-TTTCCTCCTCTCCTGACTACTCC, 604 bp), Tie2 wt (for 5’-TCCTTGCCGCCAACTTGTAAAC and rev 5’-AGCAAGCTGACTCCACAGAGAAC, 175 bp), and general Cre allele (for 5-ATGTCCAATTTACTGACCG and rev 5’-CGCCGCA TAACCAGTGAA, 673 bp). Knockout was induced with 3 doses of tamoxifen (2 mg) in peanut oil by oral gavage at 4 weeks of age. Mice for other experiment came from in house breeding on a C57BL6/J background. All experiments were performed in both female and male mice.

### Tail bleeding assay

Mice with isoflurane anesthesia were subjected to surgical dissection of the tail (3 mm from the tip). The tail was prewarmed for 2 minutes before dissection and immediately after immersed in buffered saline prewarmed to 37°C. The time of bleeding was recorded. The tail bleeding assay were performed in 6-12-week-old *Angpt2*^iECKO^ *Tie2*^iECKO^, and control mice (WT).

In addition, the same experiment was performed in WT mice 15 minutes after receiving an i.p. injection of a recombinant human His tagged fragments of ANGPT2 (ab220589, Abcam), ANGPT1 (ab69492, Abcam), Albumin (ab217817, Abcam), or IgG (ab219660, Abcam). The proteins were diluted in PBS and injected at 25 µg/kg and 250 µg/kg body weight. After the assay, heart puncture was performed to collect blood diluted 1:10 in citate-dextrose anticoagulant (C3821, Sigma), centrifuged and prepared as above. Lungs were harvested, snap frozen and stored at -80°C for later protein analysis.

### ANGPT2 association with thrombomodulin in vivo

Plasma concentrations of injected recombinant human ANGPT2 was measured by ELISA as above (DANG20, R&D Systems). Immunoprecipitation experiments were performed to evaluate the binding of ANGPT2 and ANGPT1 to thrombomodulin after injection. Lung tissue was homogenized in RIPA buffer (89901, Pierce) with proteas and phosphatase inhibitor (A32959, Pierce). Lung lysates were immunoprecipitated with a rabbit anti-mouse thrombomodulin antibody (ab230010, Abcam) attached to protein G conjugated Dynabeads (10004D, Thermo Fisher Scientific). Immunoprecipitated proteins were separated on 4-20% Mini Protean TGX gels (4561094, Biorad) and then transferred using Trans-blot turbo 0.2 µm PVDF membranes (1704156, Biorad). Blots were blocked with 5% BSA for 1 h and incubated overnight with mouse anti-6X His tag antibody (27E8, 2366, Cell Signaling). After washing and incubating with anti-mouse IgG-HRP conjugated secondary antibody (NA931, Sigma), proteins were visualized using ECL plus detection reagents (GERPN2232, Sigma). Blots were stripped with Re-Blot Plus Strong solution (2504, Millipore), blocked, and probed with anti-thrombomodulin antibody followed by anti-rabbit HRP conjugated secondary antibody (711-035-152, Jackson Immuno Research). Band density was quantified with ImageJ (NIH). Immunoprecipitation of thrombomodulin from control and patient plasma was performed as above with a rabbit anti-human thrombomodulin antibody (ab108189, Abcam) attached to protein G conjugated Dynabeads in 200 µl of plasma. ANGPT2 signal was detected with rabbit anti-human ANGPT2 (ab155106, Abcam) and donkey anti-rabbit IgG-HRP conjugated secondary antibody (711-035-152, Jackson Immuno Research). After stripping, the blot was probed with anti-human thrombomodulin antibody followed by anti-rabbit HRP conjugated secondary antibody as above. An ANGPT2 fragment (26 kDa, Ab220589, Abcam) was used to assure specificity of the ANGPT2 antibody in the experiments. All blots showed band sizes predicted by the manufacturers. Dynabeads without conjugated thrombomodulin antibody was used as negative control.

### Effect of ANGPT on thrombomodulin dependent anticoagulation

TEG on kaolin activated plasma was utilized to evaluate the effect of ANGPT2 on thrombomodulin dependent anticoagulation. Freshly drawn human citrate plasma, from both female and male donors, was incubated in kaolin tubes with 1000 ng/ml recombinant thrombomodulin (3947-PA-010, R&D Systems) and 1000 ng/ml ANGPT2 (ab220589, Abcam) at room temperature for 20 minutes. Analysis was carried out on a TEG5000 (Nordic Biolabs) for 15 minutes + reaction time (R). The measurements were started by the addition of plasma to TEG cups containing 20 µl 0.2 M CaCl_2_. Samples were run in duplicate and averaged. A sample without additives was run at the start and end of the experiment and averaged as control.

### Effect of ANGPT2 on thrombomodulin dependent activation of protein C

Pooled plasma from healthy controls was incubated with the same volume of 6 mM CaCl_2_with 0.2 U/ml thrombin (T8885, Sigma), 1000 ng/ml of recombinant thrombomodulin (3947-PA-010, R&D Systems), ANGPT2 (ab220589, Abcam), ANGPT1 (ab69492, Abcam), IgG (ab219660, Abcam) at 37°C for 30 minutes. The reaction was terminated by addition of 0.2 U/ml hirudin (H0393, Sigma) at 37°C for 10 minutes. The generation of APC was evaluated by adding chromogenic APC substrate. After addition of 50 µl chromogenic APC substrate (229021, Biophen CS-21(66)) the increase in absorbance was measured at 405 nm for 8 minutes (linear phase) at 37°C in a temperature-controlled plate reader (Synergy HT, Biotek). The area under the curve was used to calculate APC concentrations expressed as arbitrary units (a.u.).

### Study approval

The study was approved by the Swedish National Ethical Review Agency (EPM; No. 2020-01623). Informed consent was obtained from the patient, or next of kin if the patient was unable to give consent. Healthy blood donor samples were approved under ethical permit No 01/367. The Declaration of Helsinki and its subsequent revisions were followed. The protocol for the study was registered (ClinicalTrials ID: NCT04316884); STROBE guidelines were followed for reporting.

All animal experiments were approved by the Uppsala Committee of Ethics of Animal Experiments (approved permit number 5.8.18-04862-2020 and 5.8.18-03858-2021) and were conducted according to guidelines established by the Swedish Board of Agriculture.

### Statistics

Data are expressed as geometric mean ± geometric 95% CI. To test for statistical differences, we utilized Student’s *t*-test or ANOVA (>2 groups) where appropriate. ANOVA was followed by Bonferroni’s *post hoc* test. Data was unevenly normal distributed, and data was log transformed before statistical analysis. Pearson’s correlation was used to measure dependence between two variables except for SAPS-3, SOFA score, and TEG MA where non-parametric Spearman’s correlation was used. Fisher’s exact test was used to evaluate patients’ results in comparison to a given reference interval representing the normal range based on mean ± 2SD (as calculated in the clinic). Kaplan-Meier plots with Log-rank test were used to estimate the probability of survival. The optimal cut-off value was determined from Receiver Operating Characteristics (ROC) curve analysis, and the cut-off with the highest likelihood ratio was selected i.e. for ANGPT2, area under the ROC curve p=0.0004; cut-off 8.3 ng/ml with likelihood ratio 23.5; VWF, no significant area under the ROC curve.

All statistical analysis was done in GraphPad Prism 8. All analyses were 2-sided and a p<0.05 was considered significant. *<0.05, **<0.01, ***<0.001, and ****<0.0001.

## Data Availability

All data are presented within the manuscript

## Author contributions

Designing research studies: MH, MJ

Conducting experiments: MH, KF, AL, MJ

Acquiring data: MH, KF, AL, RF, ML, MJ

Analyzing data: KF, MJ

Providing reagents: SEQ, CB, MJ

Writing the manuscript: MH, KF, AL, SEQ, CB, RF, ML, MJ

## Acknowledgement

We thank Jana Chmielniakova, Pia Peterson, and Cecilia Olsson at Uppsala University for technical assistance, as well as research nurses Joanna Wessbergh and Elin Söderman, and the biobank assistants Labolina Spång, Erik Danielsson and Philip Karlsson for their expertise in compiling patient samples. We thank Peetra Magnusson at Uppsala University for valuable comments on the manuscript.

## Data and materials availability

All data are presented within the paper, individual level data can be made available on reasonable request (https://doi.org/10.17044/scilifelab.14229410).

---

**Suppl. Figure 1.**
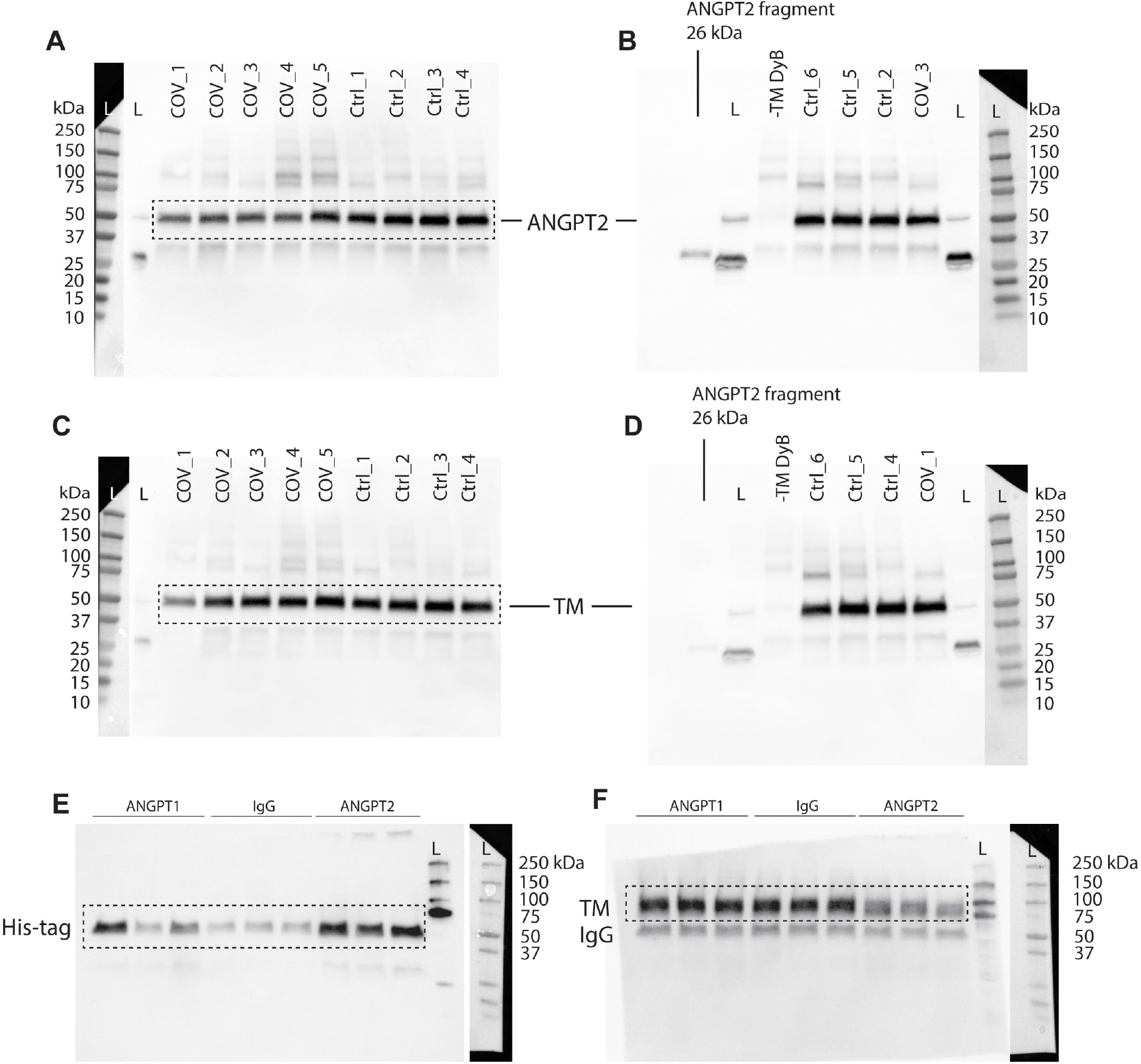
Uncut blot images. (A, B) Blots for ANGPT2 used for quantification in Fig. 1J, the dashed line marks the cut-out for Fig. 1I. (C, D) Blots for thrombomodulin used for quantification in Fig. 1J, the dashed line marks the cut-out for Fig. 1I. The pasted ladder (far left in A, C; far right in B, D) is the calorimetric signal for the ladder (L) in the blot. (E, F) Blots for His-tag (ANGPT2) and thrombomodulin, respectively, used for quantification in Fig. 2D, the dashed line marks the cut-out for Fig. 2C. The pasted ladder (far left in E, F) is the calorimetric signal for the ladder (L) in the blot.

**Suppl. Table 1.**
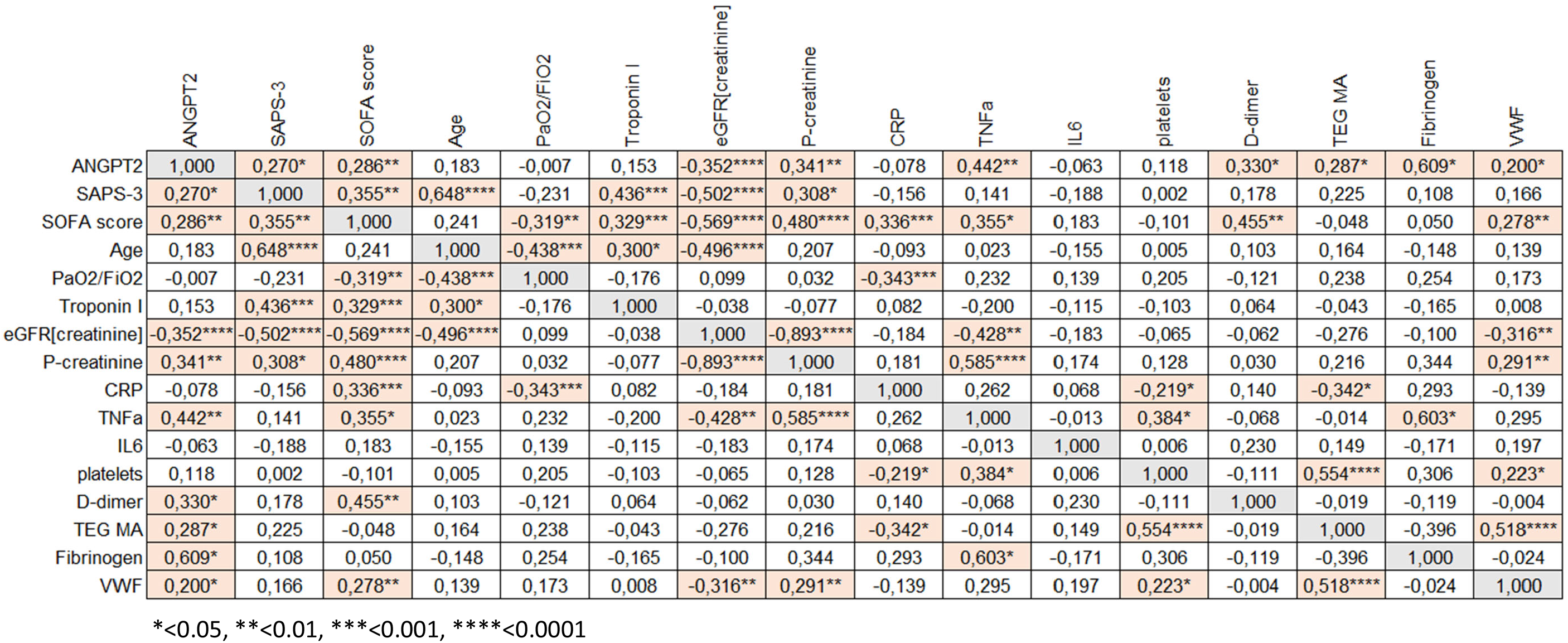
Correlations between various parameters within the cohort

